# Diencephalic or hippocampal amnesia – different etiologies, common mechanisms

**DOI:** 10.1101/2022.10.28.22281661

**Authors:** Shailendra Segobin, Melanie Ambler, Alice Laniepce, Hervé Platel, Gael Chételat, Mathilde Groussard, Anne-Lise Pitel

## Abstract

**Objectives:** To compare regional volume deficits within the Papez circuit in Alzheimer’s disease (AD) and Korsakoff’s syndrome (KS), taking into account the neurodegenerative nature of AD.

**Methods:** 18 KS patients, 40 AD patients (20 with Moderate AD (MAD) matched on cognitive deterioration with KS patients and 20 with Severe AD (SAD)), and 70 healthy controls underwent structural MRI examination. Volumes of the hippocampi, thalami, cingulate gyri, mammillary bodies (MB) and mammillothalamic tracts (MTT) were extracted.

**Results:** For the anterior and posterior cingulate gyri, and anterior thalamic nuclei, all patient groups were significantly affected compared to controls but did not differ between each other. Lower volumes were observed in all patient groups compared to controls in the entire thalamus, mediodorsal thalamic nuclei and MB, but these regions were more severely damaged in KS compared to AD. MTT volumes were significantly damaged in KS only. Hippocampi were affected in all patient groups but more severely in the SAD than in the KS and MAD.

**Interpretation:** The specificity of KS compared to AD relies on the severity of the MB and mediodorsal nuclei shrinkage, as well as the atrophy of the MTT. Several nodes of the Papez circuit were damaged to the same extent in AD and KS: the anterior thalamic nuclei, the cingulate cortex and the hippocampus (in MAD only). Our results encourage considering common mechanisms in the pathophysiology of amnesia regardless of etiology and question the relevance of the classical distinction between hippocampal and diencephalic amnesia.

## INTRODUCTION

Amnesia refers to severe episodic memory deficits that interfere with independent daily living. While amnesic cases have been described more than one century ago and have largely contributed to the understanding of memory function and substrates^1^, pathophysiology of amnesia remains unclear. Amnesic patients are rare and their neuroimaging investigation even more. However, the study of brain abnormalities in amnesic patients from different etiologies makes possible inference regarding the brain mechanisms underlying amnesia and more generally concerning the substrates of episodic memory.

Alzheimer’s disease (AD) and Korsakoff’s syndrome (KS) are two major neurocognitive disorders^2^, which both result in severe episodic memory impairments associated with loss of autonomy in daily life. AD is a neurodegenerative disease that has historically been regarded as a medial temporal lobe amnesia, with the pathology centered on hippocampal atrophy at the early stage and progressively extending to neocortical areas^3^. Contrary to AD, KS has been studied as a model of diencephalic amnesia resulting most commonly from the combination of alcohol use disorder and thiamine deficiency, and characterized by brain abnormalities especially affecting the thalami and mammillary bodies^4,5^. Post-mortem and in vivo neuroimaging investigations highlighted the key role of the anterior thalamus in the pathophysiology of KS^6,7^. However, some literature reports thalamic abnormalities in AD^8^ with neurodegeneration in the anterior thalami and a particular vulnerability of these nuclei even prodromal AD^9^. In the same vain, hippocampal volume deficits has been described in a group of five KS patients^10^ and the severity of the memory impairment correlated with the hippocampal shrinkage.

The regions crucial to both pathologies are actually part of a single brain circuit: the Papez circuit. There is increasing support in the literature to consider amnesia as the result of an impairment of a brain network responsible for memory, rather than to a particular region^11–13^. The anatomical differences and similarities between AD and KS patients can provide novel insights regarding the brain substrates of amnesia and more generally of episodic memory. In that perspective, structural abnormalities across medial temporal lobe and diencephalic brain areas need to be directly compared. As the neurodegenerative nature of AD could affect these comparison results, AD progression must be considered.

The goal of the present study is thus to compare structural brain damage in these two amnesias supposed to result from different pathophysiological mechanisms primarily involving either the hippocampus (in AD), the thalamus (in KS), or the Papez circuit in general. The present investigation therefore focuses on evaluating the volumes of regions belonging to the Papez circuit and considered hallmarks to each pathology, taking the potential effect of AD evolution into account.

## METHODS

### Population

Forty patients with AD (20 with Moderate AD (MAD) and 20 with Severe AD (SAD)), 18 patients with KS and 70 healthy controls (HC) were enrolled in this study. All groups were matched for gender and education but not for age, KS patients being younger than AD (**Table 1**). All participants (and caregivers for patients when appropriate) provided written informed consent for inclusion in the study, which was approved by the local ethics committee of Caen University Hospital (CPP Nord Ouest III, N° ID RCB: 2011-A00495-36 ; ID-RCB n°2011-A00351-40; N° ID-RCB : 2011-A01493-38 and N° ID RCB : 2007-A00414-49) in line with the Declaration of Helsinki (1964).

**Table 1:** Description of the population (mean ± standard deviation and range)

|  | HC<br>N=70 | MAD<br>patients<br>N=20 | SAD patients<br>N=20 | KS patients<br>N=18 | Statistics | Post-hoc comparisons |
| --- | --- | --- | --- | --- | --- | --- |
| **Gender (men/women)** | 24/46 | 8/12 | 2/18 | 8/10 | $\chi^2=6.36$<br>$p=0.09$ | n.a |
| **Age** | 68.3 ± 10.1<br>(50-86) | 78.7 ± 4.77<br>(70-88) | 79.5 ± 5.78<br>(71-91) | 55.6 ± 5.59<br>(44-67) | F=77.13<br>$p<0.001$ | HC<KS<(MAD=SAD) |
| **Education** | 10.1 ± 1.93<br>(6-17) | 10.1 ± 3.15<br>(6-17) | 9.05 ± 2.48<br>(6-15) | 10.1 ± 2.37<br>(6-15) | F=1.06<br>$p=0.38$ | n.a |
| **MMSE: Total Score**<br>(Max. 30) | 28.9 ± 1.10<br>(26-30) | 22.6 ± 3.03<br>(18-27) | 11.8 ± 3.56<br>(3-17) | 23.2 ± 2.71<br>(18-27) | F=185.14<br>$p<0.001$ | HC>(KS=MAD)>SAD |
| **MMSE: Orientation\***<br>(Max. 10) | 9.93± 0.315<br>(8-10) | 7.50 ± 1.46<br>(5-10) | 2.45 ± 2.04<br>(0-8) | 7.83 ± 1.72<br>(4-10) | F=204<br>$P<0.001$ | HC > (KS =MAD) > SAD |
| **MMSE: Learning\***<br>(Max. 3) | 3 ± 0.0<br>(3-3) | 2.94 ± 0.250<br>(2-3) | 2.40 ± 0.883<br>(0-3) | 2.89 ± 0.471<br>(1-3) | F=11.4<br>$p<0.001$ | (HC= KS=MAD) > SAD |
| **MMSE: Recall\***<br>(Max. 3) | 2.60 ± 0.715<br>(0-3) | 0.267 ± 0.458<br>(0-1) | 0.150 ± 0.489<br>(0-2) | 0.667 ± 0.767<br>(0-2) | F=118<br>$p<0.001$ | HC > (KS = MAD=SAD) |
| **MMSE: Attention\***<br>(Max. 5) | 4.85 ± 0.357<br>(4-5) | 3.87 ± 1.77<br>(0-5) | 0.70 ± 1.17<br>(0-4) | 3.33± 1.41<br>(0-5) | F=94.4<br>$p<0.001$ | HC>(KS=MAD)>SAD |
| **MMSE: Language\***<br>(Max. 8) | 7.56 ± 0.608<br>(5-8) | 7.07 ± 0.704<br>(6-8) | 5.85 ± 1.42<br>(1-8) | 7.50 ± 0.707<br>(6-8) | F=22.8<br>$p<0.001$ | HC=KS; HC > (MAD =SAD)<br>KS=MAD > SAD |
| **MMSE: Praxis\***<br>(Max. 1) | 0.971 ± 0.170<br>(0-1) | 0.733± 0.458<br>(0-1) | 0.300 ± 0.470<br>(0-1) | 0.722 ± 0.461<br>(0-1) | F=22.0<br>$p<0.001$ | (HC=KS=MAD) >SAD |

HC: healthy controls; MAD: moderate Alzheimer disease; SAD: severe Alzheimer disease; KS: Korsakoff's syndrome; MMSE: Mini-Mental State Examination
Post-hoc tests: Tukey HSD (unequal N); n.a = not applicable
\* MMSE subscores were missing for 2 HC and 4 MAD patients

#### Inclusion for all patients

All patients spoke French as their native language. They did not present previous neurological, psychiatric or history of severe brain injury (except brain abnormalities associated with AD or KS). No participants presented with contraindications for an MRI scan (claustrophobia, pacemaker, foreign metallic object). Clinical neuroimaging examinations ruled out other etiologies that could explain memory impairments in the patient groups.

#### Inclusion criteria specific to KS

KS patients were recruited as inpatients at Caen University Hospital (n = 9) and from a nursing home (Maison Vauban, Roubaix, France; n = 9). They met the criteria for alcohol-induced major neurocognitive disorder, amnestic-confabulatory type, persistent (DSM-5^2^). All patients presented with a history of chronic and heavy alcohol drinking even though it was difficult to obtain an accurate quantification of their alcohol consumption because of amnesia. A multidisciplinary team of specialists examined each patient to ensure an accurate diagnosis of KS. All patients had a Mini-Mental State Examination (MMSE^14^) score ≥ 18.

#### Inclusion criteria specific to AD

AD patients were recruited from the local memory clinic (Caen University Hospital) and in partnership with independent living facilities in the region of Normandy, France. All patients fulfilled the standard criteria for AD diagnosis reported by the National Institute of Neurological and Communicative Disorders and Stroke and the Alzheimer’s Disease and Related Disorders Association (NINCDS-ADRDA)^15^. Each patient was classified either in the Moderate AD (MAD) or Severe AD (SAD) group based on the results of the MMSE. MAD group included AD patients with a MMSE score ≥ 18 to match the global cognitive deterioration of KS patients. To take account of AD progression, AD patients with an MMSE score < 18 were included in the SAD group.

#### Inclusion criteria specific to HC

All HC presented age and education level appropriate performance on the MMSE. None consumed more than 3 standard drinks (2 for women) per day on a regular basis (recommendation from the World Health Organization).

#### Global cognitive description

As expected, all patient groups had lower Total MMSE score than HC, with KS and MAD presenting similar global cognitive deterioration and SAD exhibiting the most severe one. On the Recall subtest of the MMSE, the three patient groups had significantly lower results than HC but performed similarly. For the Orientation, Learning, Attention, Language and Praxis subtests of the MMSE, KS and MAD did not differ between each other but performed significantly better than SAD (except for language: MAD= SAD; **Table 1**).

### Neuroimaging examination

A high-resolution T1-weighted anatomical image was acquired for each participant on a Philips Achieva 3T scanner (Cyceron Imaging Center, Caen, France) using a 3D fast-field echo sequence (180 sagittal slices; thickness = 1 mm; repetition time = 20 ms; echo time = 4.6 ms; flip angle = 10°; field of view, 256 × 256 mm^2^; matrix, 256 × 256). Imaging data were preprocessed and analyzed using the SPM12 toolbox (www.fil.ion.ucl.ac.uk/spm). The data were normalized to the Montreal Neurological Institute (MNI) template and segmented into gray matter (GM), white matter (WM) and cerebrospinal fluid (CSF). Normalized unmodulated images from controls were averaged with a threshold at 0.5 to create a binary gray matter mask for statistical analyses. For each participant, the total intracranial volume (TIV) was calculated based on the sum of the individual volumes of GM, WM and CSF.

### Statistical analyses

Volumes from regions belonging to the Papez circuit (**Figure 1**), and considered to be the hallmarks for amnesia, were extracted for further statistical analyses. The hippocampi, thalami as well as anterior and posterior cingulate gyri, were extracted from the Hammers’ Atlas of the medial temporal lobe^16^. The anterior thalamic nuclei, the mediodorsal nuclei and mammillothalamic tracts (MTT) were separated using the thalamic histological atlas^17^. Finally, the mammillary bodies were extracted from a single-subject brain atlas^18^. Modulated gray matter maps were used to extract the volumes for all the above regions, except for the MTT, for which modulated white matter maps were used. The volumes of these ROIs were compared across groups using a generalized linear model (GLM) with the group as the independent variable (four groups: HC, MAD, SAD, KS) and age and TIV as covariates. Post-hoc tests (Tukey’s test) were conducted when appropriate.

**FIGURE 1:**
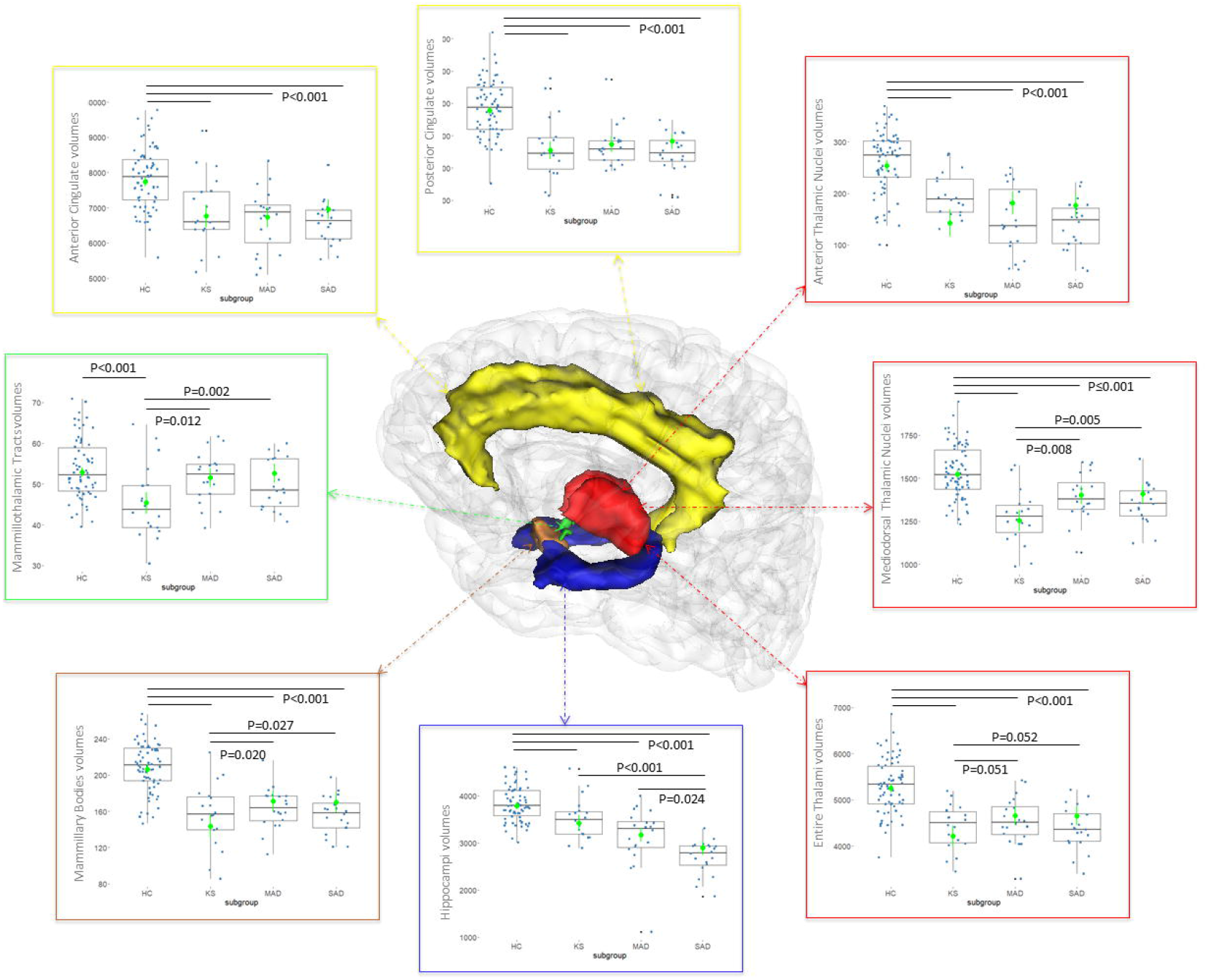
REGIONS OF THE PAPEZ CIRCUIT CONSIDERED IN THE ANALYSIS AND VOLUMETRIC COMPARISONS BETWEEN HEALTHY CONTROLS (HC), PATIENTS WITH KORSAKOFF’S SYNDROME (KS), MODERATE ALZHEIMER DISEASE (MAD) AND SEVERE ALZHEIMER DISEASE (SAD). Yellow: anterior and posterior cingulate gyri. Red: Thalamus. Blue: Hippocampus. Brown: Mammillary bodies. Green: Mammillothalamic tract. GLM: Estimated marginal means are shown in green with vertical lines showing 95% Confidence Intervals. When F-tests were significant (p<0.001, corrected for multiple comparisons), post-hoc tests were carried out (Tukey, unequal sample size). Statistically significant post-hoc tests between groups are shown with an overhead horizontal black line and associated p-value shown on the plots. Raw data are also illustrated via boxplots and volumes are expressed in mm^3^.

### Data availability

De-identified data supporting the findings of the study can be made available upon request from the corresponding author.

## RESULTS

The GLM showed that, for all 8 regions, there was a significant group effect on the regional volumes (Hippocampi F_(3,122)_=47.3; Entire thalami F_(3,122)_=38.3_;_ Anterior thalamic nuclei F_(3,122)_=38.28; Mediodorsal thalamic nuclei F_(3,122)_=30.44; Anterior cingulate gyrus F_(3,122)_=27.0; Posterior cingulate gyrus F_(3,122)_=50.6; Mammillary bodies F_(3,122)_=46.6; MTT F_(3,122)_=9.78; p<0.001 in all cases).

Further post-hoc analyses showed that the volumes of the entire thalami were significantly lower in all patient groups compared to HC (p<0.001 in all cases). Shrinkage tended to be more severe in the KS group compared to both the MAD (p=0.051) and SAD (p=0.052) groups, which did not differ from each other (p=1). The same results were observed for the mediodorsal thalamic nuclei and mammillary bodies, even though for these regions, the volumes were significantly lower in the KS group compared to both MAD (p=0.008 and p=0.02 respectively) and SAD (p=0.005 and p=0.027 respectively) groups.

The anterior thalamic nuclei, as well as the anterior and posterior cingulate gyri, were affected to the same extent in the three patient groups compared to HC (p<0.001 in all cases).

The volume of the hippocampi was significantly lower in all patient groups compared to HC (p<0.001 in all cases). Shrinkage was significantly more severe in the SAD group compared to both KS (p<0.001) and MAD (p=0.024) groups.

Finally, the volume of the MTT was lowest in the KS group and significantly different from HC (p<0.001), MAD (p=0.012) and SAD (p=0.002), who did not differ significantly from each other.

An outlier has been observed in the regional volumetric measurements in the MAD group, whose value is even lower than those from the SAD group for certain regions. Statistical analyses were performed with and without this subject and all results remained the same, bearing the same significance, albeit different p-values. Since groups of SAD and MAD are selected based on established criterion, and that there were no clinical reasons to exclude this patient, the data was kept in the analysis for the sake of completeness and integrity.

Results accounting for the two covariates are depicted in (**Figure 1)** in terms of estimated marginal means and 95% intervals. Raw data is also presented in the boxplots.

## DISCUSSION

In the existing literature, critical sites for amnesia are often reported within separate cohorts of patients, providing little opportunity to directly compare different populations experiencing amnesia, such as AD and KS. The novelty of the present study is to address this issue head-on. Data were collected from two cohorts of patients, both of which have major neurocognitive disorders characterized by amnesia, at the same site and using the same image acquisition protocols with the same MRI machine. Two groups of patients with different severities of AD were included to examine the potential effect of AD progression on the KS vs. AD comparison results.

The present study indicates that several nodes of the Papez circuit were damaged in both AD and KS amnesia, while others seem to be differentially involved in the pathophysiological mechanisms underlying these diseases. The cingulate cortex, the anterior thalamic nuclei and the hippocampus (at a moderate stage only) were damaged to the same extent in AD and KS amnesia. These two major neurocognitive disorders thus share, within the Papez circuit, a pattern of regional brain shrinkage, which could reflect the involvement of some common pathways leading to amnesia, irrespective of the etiology.

Regarding the cingulate cortex, abnormalities have previously been observed in PET studies measuring cerebral glucose metabolism in patients with severe episodic memory deficits. Hypometabolism of the posterior cingulate cortex is classically found at an early stage of AD^19^ and the cingulate cortex was shown as the only brain structure with hypometabolism in each of the 9 KS patients in a previous study^20^. Structural and functional abnormalities in the cingulate cortex are thus shared by patients with AD or KS and could be relevant to consider in the pathophysiology of these memory diseases^9,21^.

The anterior thalamic nuclei are known to play a key role in memory^21^ and their damage is considered a cardinal feature of KS^22^. As expected, we found volume deficits of the anterior thalamus in KS patients compared with controls, in accordance with previous postmortem^6^ and neuroimaging studies^23,24^. In agreement with previous studies conducted in AD^9,25,26^, we also found thalamic shrinkage in both MAD and SAD patients. Previous work described thalamus shrinkage even in patients with amnestic Mild Cognitive Impairment^8,27–29^, implicating thalamic atrophy at the early stages of the disease and in the pathophysiology of the associated episodic memory deficits. The thalamus seems especially vulnerable in prodromal AD, challenging the idea that it would only reflect a consequence of medial temporal lobe dysfunction^9^. This is in accordance with the historical description of neuropathology in the hippocampus and anterodorsal thalamus at the same stage of AD^3^. Thalamic abnormalities may thus contribute to the development of cognitive deficits in AD^30^. In theory, anterior thalamic changes would be associated with episodic memory deficits while mediodorsal thalamic changes could explain prodromal cases with predominantly executive deficits^9^.

The direct comparison of thalamic abnormalities in AD and KS provides new evidence regarding the contribution of the thalamus in amnesia. After controlling for age, AD and KS patients did not differ on the volume of the anterior thalamic nuclei, questioning the relevance of the classic nosography between hippocampal and diencephalic amnesia. In accordance with the neuropsychological profile of KS patients, frequently characterized by attention, working memory and executive dysfunction, the shrinkage of the mediodorsal nuclei^31^ was more severe in KS than in the AD groups. In patients with severe AD, such cognitive deficits may rather be related to neocortical damage.

Mammillary bodies were also affected in the three patient groups but more severely in KS than in AD. Shrinkage of the mammillary bodies is consistently described in KS^24,32,33^ but its specific contribution to the cognitive and brain pathophysiology remains unclear, mainly because it is difficult to study the impact of mamillary bodies abnormalities in isolation^5^. From an anatomical perspective, it is unlikely that mamillary body lesions explain KS amnesia since they do not affect the fornical afferents to the anterior thalamic nuclei nor the efferents from the anterior thalamic nuclei to the cingulate cortex. This leads to only a partial disconnection, which is contrary to lesions of the anterior thalamic nuclei that result in complete disconnection within the Papez circuit^5^). Mamillary body shrinkage has also been reported in AD with up to 25% of volume loss^34^, but not in patients with Mild Cognitive Impairment^35^. These findings reinforce the assumption that mamillary body abnormalities occur in patients with amnesia, potentially because of a disconnection or dysfunction in the Papez circuit, but do not initiate the pathophysiological mechanisms. The severity of the volume deficits found in KS compared with AD corroborates the hypothesis of a great vulnerability of this brain region to thiamine deficiency, leading to Wernicke’s encephalopathy and potentially to KS^5^.

The hippocampus is considered the predominant brain region responsible for episodic memory function^36^ and remains the main focus of attention in AD research. In agreement with numerous previous investigations^37,38^ and as expected, we found hippocampal shrinkage in the two groups of AD, with worsening of hippocampal atrophy with the advancement of the disease. Our findings also agree with those highlighting the hippocampus as being significantly damaged in KS patients^10,23^. To our knowledge, only one study has directly compared regional brain volume deficits in 5 KS patients versus in 20 AD patients^10^. After controlling for age differences between groups, the authors found comparable hippocampal volume deficits, which related to memory impairments in both groups. We found similar results in the present study which included a much larger group of KS and with groups of MAD and KS matched for memory performance and global cognitive deterioration. Altogether, these findings further encourage considering the notion of common mechanisms in the pathophysiology of amnesia, irrespective of the etiology.

The MTT were observed as damaged in the KS group only, suggesting that its alterations could be specific to diencephalic amnesia. The MTT effectively connects the mammillary bodies, which itself receives input from the hippocampus, to the anterior thalamic nuclei (ATN)^39^. Lesions to the MTT bilaterally, accompanied by damage to the ATN, have been sufficient to cause acute and irreversible memory disorder that is very similar to alcoholic KS^40^. Furthermore, another study^41^ showed that 7 out of 12 patients with thalamic infarct who had damage to the MTT performed worse in verbal episodic memory task than the 5 patients with preserved MTT. Taken together, these suggest that lesions to the anterior thalamic nuclei could be, to a certain extent, the result of a disconnection process through the MTT, having failed to receive part of the inputs from the hippocampus coming through the mammillary bodies^39,42^.

In conclusion, in a study which includes groups of patients that are difficult and rare to examine from a research point of view, our findings provide new direct evidence regarding the specificities and commonalities in the pattern of volume deficits within the Papez circuit in AD and KS, two major neurocognitive disorders leading to amnesia. Our study stems into the broader consideration of the substrates of amnesia and challenges the relevance of considering amnesia in reference to classic descriptions as the result of critical damage to one non-overlapping region rather than the aftermath of a cascade of pathophysiological events within the Papez circuit leading to a neuropsychological syndrome. From a theoretical perspective, these data reinforce the relevance of examining not only the hippocampus or the thalamus, but a network of connected brain regions as the substrate of memory functioning^21^.

## Acknowledgment

The authors would like to thank the “Hom’Age group”, Odile Letortu and Vincent de la Sayette for their help in the recruitment of AD patients, as well as the “Maison Vauban” (Laurent Urso) and Nicolas Cabé for their help in the recruitment of KS patients. We are also grateful to the neuropsychologists involved in this study (Caroline Mauger, Marion Hommet, Céline Boudehent and Coralie Lannuzel) and other collaborators (François Vabret, Hélène Beaunieux, Ludivine Ritz, Arnaud Mortier).

## Contribution of each author to the study

Conception and design of the study: SS, MA, MG, ALP

Acquisition and analysis of data: SS, MA, AL, MG, ALP

Drafting a significant portion of the manuscript or figures: SS, MA, AL, GC, HP, MG, ALP

## Conflict of interest

Nothing to report.

## Notes

### Competing Interest Statement

The authors have declared no competing interest.

### Funding Statement

This study did not receive any funding. One member received financial support of fulbright program to study in France on this project

### Author Declarations

Local ethics committee of Caen University Hospital gave ethical approval for this work (CPP Nord Ouest III, N ID RCB: 2011-A00495-36 ; ID-RCB n2011-A00351-40; N ID-RCB : 2011-A01493-38 and N ID RCB : 2007-A00414-49)

## REFERENCES

1. WB S, B M. Loss of recent memory after bilateral hippocampal lesions. 1957. J. Neuropsychiatry Clin. Neurosci. 2000;12(1):103–103.

2. American Psychiatric Association. Diagnostic and Statistical Manual of mental disorders: DSM-5. American P. Arlington: 2013.

3. Braak H, Braak E. Neuropathological stageing of Alzheimer-related changes. Acta Neuropathol. 1991;82(4):239–259.

4. Segobin S, Pitel AL. The specificity of thalamic alterations in Korsakoff’s syndrome: Implications for the study of amnesia. Neurosci. Biobehav. Rev. 2021;130:292–300.

5. Arts NJM, Pitel AL, Kessels RPC. The contribution of mamillary body damage to Wernicke’s encephalopathy and Korsakoff’s syndrome. Handb. Clin. Neurol. 2021;180:455–475.

6. Harding A, Halliday G, Caine D, Kril J. Degeneration of anterior thalamic nuclei differentiates alcoholics with amnesia. Brain 2000;123 (Pt 1)(1):141–154.

7. Segobin S, Laniepce A, Ritz L, et al. Dissociating thalamic alterations in alcohol use disorder defines specificity of Korsakoff’s syndrome. Brain 2019;142(5):1458–1470.

8. Bernstein AS, Rapcsak SZ, Hornberger M, Saranathan M. Structural Changes in Thalamic Nuclei Across Prodromal and Clinical Alzheimer’s Disease. J. Alzheimers. Dis. 2021;82(1):361–371.

9. Aggleton JP, Pralus A, Nelson AJD, Hornberger M. Thalamic pathology and memory loss in early Alzheimer’s disease: moving the focus from the medial temporal lobe to Papez circuit. Brain 2016;aww083.

10. Sullivan E V., Marsh L. Hippocampal volume deficits in alcoholic Korsakoff’s syndrome. Neurology 2003;61(12):1716–1719.

11. Aggleton JP. Looking beyond the hippocampus: old and new neurological targets for understanding memory disorders. Proceedings. Biol. Sci. 2014;281(1786)

12. Forno G, Lladó A, Hornberger M. Going round in circles-The Papez circuit in Alzheimer’s disease. Eur. J. Neurosci. 2021;54(10):7668–7687.

13. Aggleton JP, Pralus A, Nelson AJD, Hornberger M. Thalamic pathology and memory loss in early Alzheimer’s disease: moving the focus from the medial temporal lobe to Papez circuit. Brain 2016;139(Pt 7):1877–1890.

14. Folstein MF, Folstein SE, McHugh PR. “Mini-mental state”. A practical method for grading the cognitive state of patients for the clinician. J. Psychiatr. Res. 1975;12(3):189–198.

15. McKhann GM. Changing concepts of Alzheimer disease. JAMA 2011;305(23):2458–2459.

16. Hammers A, Allom R, Koepp MJ, et al. Three-dimensional maximum probability atlas of the human brain, with particular reference to the temporal lobe. Hum. Brain Mapp. 2003;19(4):224–247.

17. Krauth A, Blanc R, Poveda A, et al. A mean three-dimensional atlas of the human thalamus: generation from multiple histological data. Neuroimage 2010;49(3):2053–2062.

18. Holmes CJ, Hoge R, Collins L, et al. Enhancement of MR images using registration for signal averaging. J. Comput. Assist. Tomogr. 1998;22(2):324–333.

19. Chételat G, Villain N, Desgranges B, et al. Posterior cingulate hypometabolism in early Alzheimer’s disease: what is the contribution of local atrophy versus disconnection? Brain 2009;132(Pt 12)

20. Pitel AL, Aupée AM, Chételat G, et al. Morphological and glucose metabolism abnormalities in alcoholic Korsakoff’s syndrome: group comparisons and individual analyses. PLoS One 2009;4(11)

21. Aggleton JP, O’Mara SM. The anterior thalamic nuclei: core components of a tripartite episodic memory system. Nat. Rev. Neurosci. 2022;

22. Pitel AL, Segobin SH, Ritz L, et al. Thalamic abnormalities are a cardinal feature of alcohol-related brain dysfunction. Neurosci. Biobehav. Rev. 2015;54:38–45.

23. Mayes AR, Meudell PR, Mann D, Pickering A. Location of lesions in Korsakoff’s syndrome: neuropsychological and neuropathological data on two patients. Cortex. 1988;24(3):367–388.

24. Pitel A-L, Chételat G, Le Berre AP, et al. Macrostructural abnormalities in Korsakoff syndrome compared with uncomplicated alcoholism. Neurology 2012;78(17):1330–1333.

25. De Jong LW, Van Der Hiele K, Veer IM, et al. Strongly reduced volumes of putamen and thalamus in Alzheimer’s disease: an MRI study. Brain 2008;131(Pt 12):3277–3285.

26. Cherubini A, Péran P, Spoletini I, et al. Combined volumetry and DTI in subcortical structures of mild cognitive impairment and Alzheimer’s disease patients. J. Alzheimers. Dis. 2010;19(4):1273–1282.

27. De Oliveira MS, Balthazar MLF, D’Abreu A, et al. MR imaging texture analysis of the corpus callosum and thalamus in amnestic mild cognitive impairment and mild Alzheimer disease. AJNR. Am. J. Neuroradiol. 2011;32(1):60–66.

28. Pedro T, Weiler M, Yasuda CL, et al. Volumetric brain changes in thalamus, corpus callosum and medial temporal structures: mild Alzheimer’s disease compared with amnestic mild cognitive impairment. Dement. Geriatr. Cogn. Disord. 2012;34(3–4):149–155.

29. Karas GB, Scheltens P, Rombouts SARB, et al. Global and local gray matter loss in mild cognitive impairment and Alzheimer’s disease. Neuroimage 2004;23(2):708–716.

30. Swartz RH, Black SE. Anterior-medial thalamic lesions in dementia: frequent, and volume dependently associated with sudden cognitive decline. J. Neurol. Neurosurg. Psychiatry 2006;77(12):1307–1312.

31. Pergola G, Danet L, Pitel AL, et al. The Regulatory Role of the Human Mediodorsal Thalamus. Trends Cogn. Sci. 2018;22(11):1011–1025.

32. Shear PK, Sullivan E V., Lane B, Pfefferbaum A. Mammillary body and cerebellar shrinkage in chronic alcoholics with and without amnesia. Alcohol. Clin. Exp. Res. 1996;20(8):1489–1495.

33. Sullivan E V., Lane B, Deshmukh A, et al. In vivo mammillary body volume deficits in amnesic and nonamnesic alcoholics. Alcohol. Clin. Exp. Res. 1999;23(10):1629–1636.

34. Sheedy D, Lara A, Garrick T, Harper C. Size of mamillary bodies in health and disease: useful measurements in neuroradiological diagnosis of Wernicke’s encephalopathy. Alcohol. Clin. Exp. Res. 1999;23(10):1624–1628.

35. Copenhaver BR, Rabin LA, Saykin AJ, et al. The fornix and mammillary bodies in older adults with Alzheimer’s disease, mild cognitive impairment, and cognitive complaints: a volumetric MRI study. Psychiatry Res. 2006;147(2–3):93–103.

36. Squire LR, Stark CEL, Clark RE. The medial temporal lobe. Annu. Rev. Neurosci. 2004;27:279–306.

37. de Flores R, La Joie R, Ch??telat G. Structural imaging of hippocampal subfields in healthy aging and Alzheimer’s disease. Neuroscience 2015;309:29–50.

38. Wei X, Du X, Xie Y, et al. Mapping cerebral atrophic trajectory from amnestic mild cognitive impairment to Alzheimer’s disease. Cereb. Cortex 2022;

39. Dillingham CM, Milczarek MM, Perry JC, Vann SD. Time to put the mammillothalamic pathway into context. Neurosci. Biobehav. Rev. 2021;121:60–74.

40. Yoneoka Y, Seki Y, Akiyama K. “Vascular” Korsakoff Syndrome With Bilaterally Damaged Mammillothalamic Tracts: Insights Into the Pathogenesis of “Acute” Korsakoff Syndrome As Acute-Onset Irreversible Anterograde Amnesia. Cureus 2021;13(11)

41. Danet L, Barbeau EJ, Eustache P, et al. Thalamic amnesia after infarct: The role of the mammillothalamic tract and mediodorsal nucleus. Neurology 2015;85(24):2107–2115.

42. Vann SD, Nelson AJD. The mammillary bodies and memory: more than a hippocampal relay. Prog. Brain Res. 2015;219:163–185.

